# Immune Responses in Fully Vaccinated Individuals Following Breakthrough Infection with the SARS-CoV-2 Delta Variant in Provincetown, Massachusetts

**DOI:** 10.1101/2021.10.18.21265113

**Authors:** Ai-ris Y. Collier, Catherine M. Brown, Katherine Mcmahan, Jingyou Yu, Jinyan Liu, Catherine Jacob-Dolan, Abishek Chandrashekar, Dylan Tierney, Jessica L. Ansel, Marjorie Rowe, Daniel Sellers, Kunza Ahmad, Ricardo Aguayo, Tochi Anioke, Sarah Gardner, Mazuba Siamatu, Lorraine Bermudez Rivera, Michele R. Hacker, Lawrence C. Madoff, Dan H. Barouch

## Abstract

**Background:** A cluster of over a thousand infections with the SARS-CoV-2 delta variant was identified in a predominantly fully vaccinated population in Provincetown, Massachusetts in July 2021. Immune responses in breakthrough infections with the SARS-CoV-2 delta variant remain to be defined.

**Methods:** Humoral and cellular immune responses were assessed in 35 vaccinated individuals who were tested for SARS-CoV-2 in the Massachusetts Department of Public Health outbreak investigation.

**Results:** Vaccinated individuals who tested positive for SARS-CoV-2 demonstrated substantially higher antibody responses than vaccinated individuals who tested negative for SARS-CoV-2, including 28-fold higher binding antibody titers and 34-fold higher neutralizing antibody titers against the SARS-CoV-2 delta variant. Vaccinated individuals who tested positive also showed 4.4-fold higher Spike-specific CD8+ T cell responses against the SARS-CoV-2 delta variant than vaccinated individuals who tested negative.

**Conclusions:** Fully vaccinated individuals developed robust anamnestic antibody and T cell responses following infection with the SARS-CoV-2 delta variant. These data suggest important immunologic benefits of vaccination in the context of breakthrough infections.

## INTRODUCTION

A large cluster of COVID-19 infections were reported to the Massachusetts Department of Public Health (MA DPH) after the July 4, 2021 weekend in Provincetown, Barnstable County, Massachusetts^1^. Approximately 74% of cases occurred in individuals who were fully vaccinated with the BNT162b2 (Pfizer/BioNTech), mRNA-1273 (Moderna), or Ad26.COV.2 (Johnson & Johnson) COVID-19 vaccines^2-4^. Most cases were mildly or moderately symptomatic. Viral sequencing revealed that over 90% of cases sequenced were the SARS-CoV-2 delta variant, and viral loads in nasal swabs were similar in vaccinated and unvaccinated individuals^1^. This outbreak was the first known large cluster of infections with the SARS-CoV-2 delta variant in a highly vaccinated population, and it prompted the U.S. Centers for Disease Control and Prevention (CDC) to reinstate indoor masking recommendations for fully vaccinated individuals.

We recruited vaccinated individuals who were part of the MA DPH outbreak investigation or enhanced surveillance and who tested positive or negative for COVID-19 by nasopharyngeal swabs to participate in a detailed immunologic study at Beth Israel Deaconess Medical Center (BIDMC) in Boston, Massachusetts. We measured peripheral and mucosal antibody responses as well as cellular immune responses in vaccinated individuals with or without SARS-CoV-2 breakthrough infection.

## METHODS

### Study population

To recruit participants who were part of the outbreak investigation of the breakthrough infections in Provincetown, Massachusetts, the MA DPH and Boston Public Health Commission (BPHC) provided information about this immunologic study to individuals by telephone or email. Participants were provided contact information for the Beth Israel Deaconess Medical Center (BIDMC) study team for recruitment and informed consent. Participants also self-referred from flyers posted on social media. Participants were asked to provide their vaccine, symptom, and testing history, as well as their race and ethnicity based on specified categories; they could select multiple race categories. The BIDMC Institutional Review Board approved this study (#2021P000344) as part of a parent biorepository study (#2020P000361). All participants provided informed consent. Breakthrough SARS-CoV-2 infection was defined as a positive nasopharyngeal swab PCR test after being fully vaccinated (at least 2 weeks following two doses of the BNT162b2 or mRNA-1273 vaccines or at least 2 weeks after a single dose of the Ad26.COV2.S vaccine). Participants were asked to provide blood samples and nasal swabs.

### Pseudovirus neutralizing antibody assay

The SARS-CoV-2 pseudoviruses expressing a luciferase reporter gene were used to measure pseudovirus neutralizing antibodies as described previously.^5,6^ In brief, the packaging construct psPAX2 (AIDS Resource and Reagent Program), luciferase reporter plasmid pLenti-CMV Puro-Luc (Addgene) and spike protein expressing pcDNA3.1-SARS-CoV-2 SΔCT were co-transfected into HEK293T cells (ATCC CRL_3216) with lipofectamine 2000 (ThermoFisher Scientific). Pseudoviruses of SARS-CoV-2 variants were generated by using WA1/2020 strain (Wuhan/WIV04/2019, GISAID accession ID: EPI_ISL_402124), B.1.1.7 variant (Alpha, GISAID accession ID: EPI_ISL_601443), B.1.351 variant (Beta, GISAID accession ID: EPI_ISL_712096), or B.1.617.2 (Delta, GISAID accession ID: EPI_ISL_2020950). The supernatants containing the pseudotype viruses were collected 48h after transfection; pseudotype viruses were purified by filtration with 0.45-μm filter. To determine the neutralization activity of human serum, HEK293T-hACE2 cells were seeded in 96-well tissue culture plates at a density of 1.75 × 10^4^ cells per well overnight. Three-fold serial dilutions of heat-inactivated serum samples were prepared and mixed with 50 μl of pseudovirus. The mixture was incubated at 37 °C for 1 h before adding to HEK293T-hACE2 cells. After 48 h, cells were lysed in Steady-Glo Luciferase Assay (Promega) according to the manufacturer’s instructions. SARS-CoV-2 neutralization titers were defined as the sample dilution at which a 50% reduction (NT50) in relative light units was observed relative to the average of the virus control wells.

### Enzyme-linked immunosorbent assay (ELISA)

SARS-CoV-2 spike receptor-binding domain (RBD)-specific binding antibodies in serum were assessed by ELISA as described previously.^5^ 96-well plates were coated with 2 μg/mL of SARS-CoV-2 WA1/2020, Alpha, Beta, Gamma, Delta, or Kappa RBD protein (source: F. Krammer, Icahn School of Medicine at Mount Sinai) in 1× Dulbecco phosphate-buffered saline (DPBS) and incubated at 4 °C overnight. After incubation, plates were washed once with wash buffer (0.05% Tween 20 in 1× DPBS) and blocked with 350 μL of casein block solution per well for 2-3 h at room temperature. Following incubation, block solution was discarded and plates were blotted dry. Serial dilutions of heat-inactivated serum diluted in Casein block or nasal swab sample diluted in DPBS were added to wells, and plates were incubated for 1 hour at room temperature, prior to 3 more washes and a 1-hour incubation with a 1:4000 dilution of anti–human IgG horseradish peroxidase (HRP) 1:1000 dilution of anti-human IgA horseradish peroxidase (Bethyl Laboratories) at room temperature in the dark. Plates were washed 3 times, and 100 μL of SeraCare KPL TMB SureBlue Start solution was added to each well; plate development was halted by adding 100 μL of SeraCare KPL TMB Stop solution per well. The absorbance at 450 nm, with a reference at 650 nm, was recorded with a VersaMax microplate reader (Molecular Devices). For each sample, the ELISA end point titer was calculated using a 4-parameter logistic curve fit to calculate the reciprocal serum dilution that yields a corrected absorbance value (450 nm-650 nm) of 0.2. Interpolated end point titers were reported.

### Electrochemiluminescence assay (ECLA)

ECLA plates (MesoScale Discovery SARS-CoV-2 IgG Cat No: N05CA-1; Panel 7 and K15463U-2; Panel 13) were designed and produced with up to 9 antigen spots in each well, and assays were performed essentially as described previously.^10^ The antigens included WA1/2020, B.1.1.7 (alpha), B.1.351 (beta), P.1 (gamma), B.1.617.2 (delta), and B.1.617.1 (kappa) and WA1/2020 Nucleocapsid (Nuc). The plates were blocked with 50 uL of Blocker A (1% BSA in distilled water) solution for at least 30 minutes at room temperature shaking at 700 rpm with a digital microplate shaker.

During blocking the serum was diluted to 1:5,000 in Diluent 100. The calibrator curve was prepared by diluting the calibrator mixture from MSD 1:9 in Diluent 100 and then preparing a 7-step 4-fold dilution series plus a blank containing only Diluent 100. The plates were then washed 3 times with 150 μL of Wash Buffer (0.5% Tween in 1x PBS), blotted dry, and 50 μL of the diluted samples and calibration curve were added in duplicate to the plates and set to shake at 700 rpm at room temperature for at least 2 h. The plates were again washed 3 times and 50 μL of SULFO-Tagged anti-Human IgG detection antibody diluted to 1x in Diluent 100 was added to each well and incubated shaking at 700 rpm at room temperature for at least 1 h. Plates were then washed 3 times and 150 μL of MSD GOLD Read Buffer B was added to each well and the plates were read immediately after on a MESO QuickPlex SQ 120 machine. MSD titers for each sample were reported as relative light units (RLU), which were calculated using the calibrator.

### Intracellular cytokine staining (ICS) assay

10^6^ peripheral blood mononuclear cells well were re-suspended in 100 µL of R10 media supplemented with CD49d monoclonal antibody (1 µg/mL) and CD28 monoclonal antibody (1 µg/mL) as described previously. Each sample was assessed with mock (100 µL of R10 plus 0.5% DMSO; background control), peptides (2 µg/mL), and/or 10 pg/mL phorbol myristate acetate (PMA) and 1 µg/mL ionomycin (Sigma-Aldrich) (100µL; positive control) and incubated at 37°C for 1 h. After incubation, 0.25 µL of GolgiStop and 0.25 µL of GolgiPlug in 50 µL of R10 was added to each well and incubated at 37°C for 8 h and then held at 4°C overnight. The next day, the cells were washed twice with DPBS, stained with aqua live/dead dye for 10 mins and then stained with predetermined titers of monoclonal antibodies against CD279 (clone EH12.1, BB700), CD4 (clone L200, BV711), CD27 (clone M-T271, BUV563), CD8 (clone SK1, BUV805), CD45RA (clone 5H9, APC H7) for 30 min. Cells were then washed twice with 2% FBS/DPBS buffer and incubated for 15 min with 200 µL of BD CytoFix/CytoPerm Fixation/Permeabilization solution. Cells were washed twice with 1X Perm Wash buffer (BD Perm/WashTM Buffer 10X in the CytoFix/CytoPerm Fixation/ Permeabilization kit diluted with MilliQ water and passed through 0.22µm filter) and stained with intracellularly with monoclonal antibodies against IFN-γ (clone B27; BUV395), and CD3 (clone SP34.2, Alexa 700), for 30 min. Cells were washed twice with 1X Perm Wash buffer and fixed with 250µL of freshly prepared 1.5% formaldehyde. Fixed cells were transferred to 96-well round bottom plate and analyzed by BD FACSymphony™ system. Data were analyzed using FlowJo v9.9.

### Enzyme-linked immunospot (ELISPOT) assay

ELISPOT plates were coated with mouse anti-human IFN-γ monoclonal antibody from MabTech at 1 µg/well and incubated overnight at 4°C. Plates were washed with DPBS, and blocked with R10 media (RPMI with 10% heat inactivated FBS with 1% of 100x penicillin-streptomycin, 1M HEPES, 100mM Sodium pyruvate, 200mM L-glutamine, and 0.1% of 55mM 2-Mercaptoethanol) for 2-4 h at 37°C. SARS-CoV-2 pooled S peptides from WA1/2020, B.1.351, B.1.1.7, and B.1.617.2 (21st Century Biochemicals) were prepared and plated at a concentration of 2 µg/well, and 100,000 cells/well were added to the plate. The peptides and cells were incubated for 15-20 h at 37°C. All steps following this incubation were performed at room temperature. The plates were washed with ELISPOT wash buffer and incubated for 2-4 h with Biotinylated mouse anti-human IFN-γ monoclonal antibody from MabTech (1 µg/mL). The plates were washed a second time and incubated for 2-3 h with conjugated goat anti-biotin AP from Rockland, Inc. (1.33 µg/mL). The final wash was followed by the addition of Nitor-blue Tetrazolium Chloride/5-bromo-4-chloro 3 ‘indolyphosphate p-toludine salt (NBT/BCIP chromagen) substrate solution for 7 min. The chromagen was discarded and the plates were washed with water and dried in a dim place for 24 h. Plates were scanned and counted on a Cellular Technologies Limited Immunospot Analyzer.

### Statistical analysis

Descriptive statistics were calculated using GraphPad Prism 8.4.3, (GraphPad Software, San Diego, California). Data are presented as median with interquartile range (IQR). Comparisons were performed using the Wilcoxon rank-sum test. Spearman correlation coefficient (r) was reported for antibody titers with time from first vaccine dose and age. All tests were two-tailed. When all six variants were compared between infected and uninfected or between BNT162b2 and mRNA-1273, a p value <0.01 was considered statistically significant to account for multiple comparisons. For all other comparisons, p values<0.05 were considered statistically significant.

## RESULTS

### Participants

A total of 35 individuals from the MA DPH outbreak investigation^1^ were enrolled, including 14 vaccinated individuals who tested positive for SARS-CoV-2 and 21 vaccinated individuals who tested negative for SARS-CoV-2 (**Table 1**). Participants were predominantly white, and the majority were male. Vaccinated infected individuals were generally younger (median age 38 years; range 25-60) than vaccinated uninfected individuals (median age 61 years; range 24-77). Participants received the BNT162b2 (Pfizer; n=13), mRNA-1273 (Moderna; n=21), or Ad26.COV2.S (Johnson & Johnson; n=1) vaccines. Peripheral blood and nasal swab samples were obtained for immunologic assays at a median of 183 days (range 127-251) following first vaccine dose and a median of 34 days (range 0-67) following nasopharyngeal SARS-CoV-2 PCR testing in the outbreak investigation. In the vaccinated infected group, 14 of 14 (100%) of individuals reported symptoms of COVID-19 infection, most commonly respiratory symptoms, fever, and loss of smell or taste, consistent with the overall outbreak investigation;^1^ all had mild disease according to the NIH disease severity classification. None required hospitalization.

**Table 1.** Characteristics of study population

|  | Vaccinated,<br>infected<br>n=14 | Vaccinated,<br>uninfected<br>n=21 |
| --- | --- | --- |
| Age (years), median (IQR) | 38 (36-40) | 61 (54-66) |
| Sex at birth, female, n (%) | 3 (21) | 6 (29) |
| Race, n (%) |  |  |
| White | 12 (86) | 20 (95) |
| Black | 0 | 0 |
| Asian | 1 (7) | 1 (5) |
| More than one race | 0 | 0 |
| Another race | 1 (7) | 0 |
| Ethnicity, n (%) |  |  |
| Hispanic or Latino | 0 | 1 (5) |
| Non-Hispanic | 14 (100) | 20 (95) |
| Medical condition, n (%) |  |  |
| Pregnant | 0 | 0 |
| Diabetes | 0 | 2 (10) |
| Hypertension | 3 (21) | 7 (33) |
| Obesity (Body mass index ≥ kg/m<sup>2</sup>) | 3 (21) | 3 (14) |
| Asthma | 3 (21) | 3 (14) |
| Immunosuppression therapy | 0 | 1 (5) |
| COVID19 Vaccine, n (%) |  |  |
| BNT162b2 | 5 (36) | 8 (38) |
| mRNA-1273 | 8 (57) | 13 (62) |
| Ad26.COV2.S | 1 (7) | 0 |
| Days from 1st vaccine dose to sampling, median (IQR) | 175 (135-212) | 189 (148-222) |
| Days from PCR test to sampling, median (IQR) | 33 (22-40) | 34 (27-38) |
| Symptoms, n | 14 | 15 |
| Never symptomatic, n (%) | 0 | 9 (60) |
| Respiratory, n (%) | 13 (93) | 5 (33) |
| Gastrointestinal, n (%) | 2 (14) | 0 |
| Fever, n (%) | 8 (57) | 0 |
| Sore throat, n (%) | 7 (50) | 1 (7) |
| Loss of smell or taste, n (%) | 13 (93) | 0 |
| Other, n (%) | 9 (64) | 5 (33) |
| NIH disease severity, n (%) |  |  |
| Asymptomatic | 0 | N/A |
| Mild/Moderate | 14 (100) | N/A |
| Severe/Critical | 0 | N/A |

IQR= interquartile range
PCR= polymerase chain reaction nasopharyngeal testing for SARS-CoV-2
NIH= National Institutes for Health

### Humoral Immune Responses

SARS-CoV-2 antibody responses in serum were assessed by Spike- and Nucleocapsid-specific electrochemiluminescence assays (ECLAs), receptor binding domain (RBD)-specific IgG enzyme-linked immunosorbent assays (ELISAs), and pseudovirus neutralizing antibody assays. Spike-specific ECLA titers against the SARS-CoV-2 WA1/2020, B.1.1.7 (alpha), B.1.351 (beta), P.1 (gamma), B.1.617.2 (delta), and B.1.617.1 (kappa) variants were 12-, 15-, 11-, 12-, 16-, and 13-fold higher, respectively, in vaccinated infected individuals compared with vaccinated uninfected individuals (**Fig. 1A**; P<0.001 for all variants, Wilcoxon rank-sum tests). Nucleocapsid-specific ECLA responses were detected in all vaccinated infected participants, consistent with SARS-CoV-2 infection, and in only one vaccinated uninfected participant (**Fig. 1B**; P<0.001, Wilcoxon rank-sum tests). RBD-specific IgG ELISA titers against the SARS-CoV-2 WA1/2020, alpha, beta, gamma, delta, and kappa variants were 20-, 20-, 30-, 20-, 28-, and 27-fold higher, respectively, in vaccinated infected individuals compared with vaccinated uninfected individuals (**Fig. 1C**; P<0.001 for all variants, Wilcoxon rank-sum tests). Pseudovirus neutralizing antibody titers against the SARS-CoV-2 WA1/2020, alpha, beta, and delta variants were 13-, 16-, 73-, and 34-fold higher, respectively, in vaccinated infected individuals compared with vaccinated uninfected individuals (**Fig. 1D**; P<0.001 for all variants, Wilcoxon rank-sum tests).

**Figure 1.**
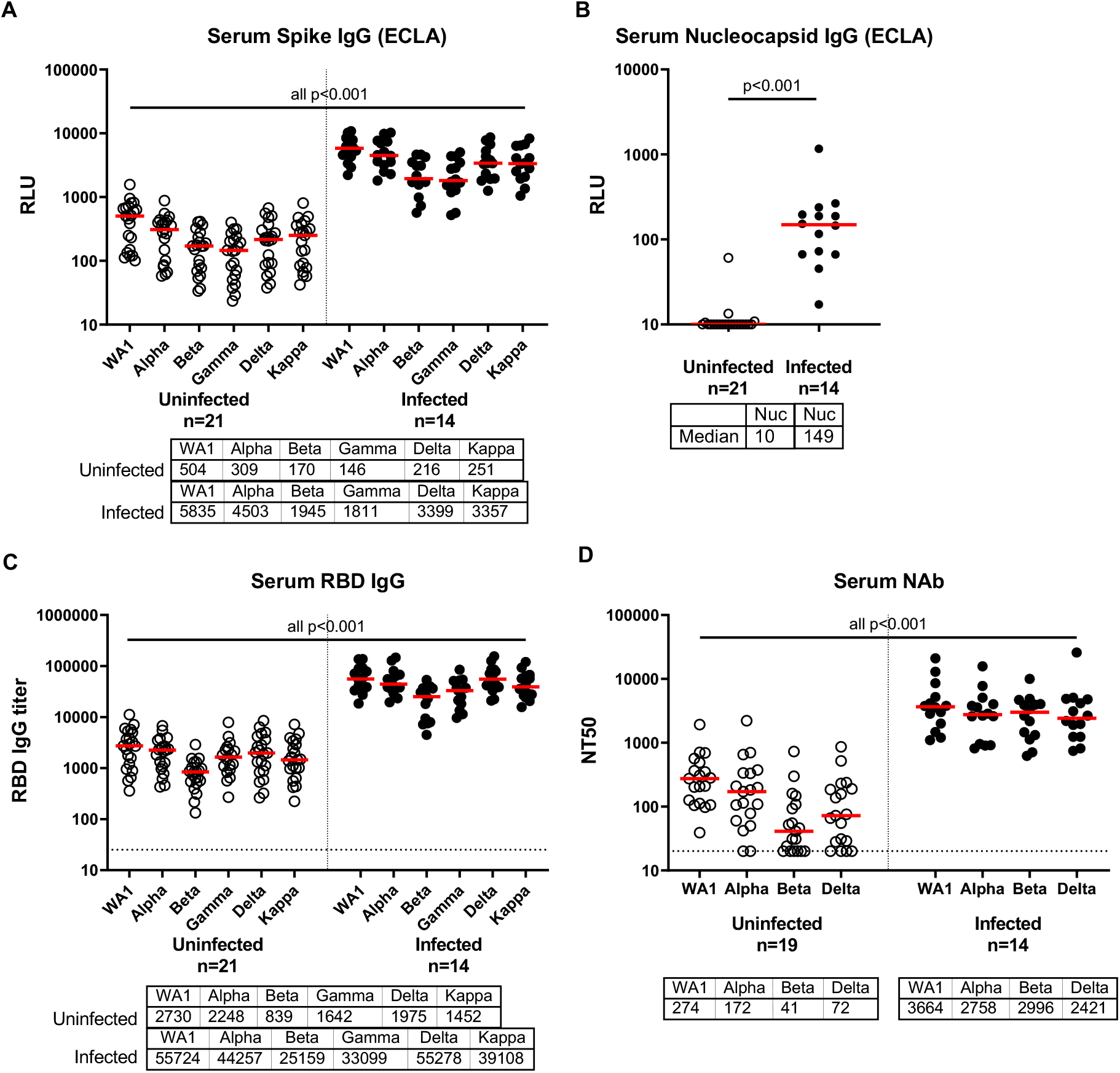
Antibody responses in COVID-19 vaccinated individuals with and without confirmed SARS-CoV-2 breakthrough infection Serum antibody responses in BNT162b2, mRNA-1273, and Ad26.COV2.S vaccinated individuals as part of the SARS-CoV-2 outbreak investigation in Provincetown, Massachusetts. Vaccinated uninfected (open circles) and vaccinated infected (filled circles) individuals are shown. IgG binding antibody titers by multiplexed electrochemiluminescence (ECLA) assays against SARS-CoV-2 Spike from WA1/2020, B.1.1.7 (alpha), B.1.351 (beta), P.1 (gamma), B.1.617.2 (delta), and B.1.617.1 (kappa) variants in **A** and against the non-vaccine antigen Nucleocapsid (Nuc) in **B** are displayed in relative light units (Meso-Scale Discovery kits 7 and 13). **C**. ELISA IgG titers to SARS-COV-2 receptor binding domain (RBD) variants. **D**. Pseudovirus neutralization titers measured as 50% reduction (NT50) of luciferase expression. Medians (red bar) are displayed below the x-axis. Dotted line indicates limit of detection. P values (Wilcoxon rank-sum test) compare each Spike variant or Nucleocapsid between uninfected and infected participants..

Similar trends were observed in subgroup analyses of participants who received BNT162b2 or mRNA-1273 (**Fig. S1**). Only one individual received Ad26.COV2.S in this study. Among vaccinated uninfected individuals, binding and neutralizing antibody responses induced by mRNA-1273 were higher than BNT162b2 responses, and the majority of individuals who received BNT162b2 had undetectable neutralizing antibody responses against the SARS-CoV-2 delta variant at the timepoint analyzed (**Fig. S1C**). No correlations were observed between participant age and the titers of binding and neutralizing antibodies in this cohort (**Fig. S2**). Moreover, robust antibody titers were observed in all vaccinated infected individuals, regardless of time from vaccination (**Fig. S3**). Taken together, these data demonstrate markedly higher binding and neutralizing antibody responses in vaccinated individuals who tested positive for SARS-CoV-2 compared with vaccinated individuals who tested negative for SARS-CoV-2.

### Cellular Immune Responses

SARS-CoV-2 cellular immune responses were assessed by intracellular cytokine staining (ICS) assays and ELISPOT assays using pooled Spike peptides. Spike-specific CD4+ T cell responses to SARS-CoV-2 WA1/2020 and B.1.617.2 (delta) were 2.1- and 1.8-fold higher, respectively, in vaccinated infected individuals compared with vaccinated uninfected individuals (**Fig. 2A**; P<0.001 and P=0.02, respectively, Wilcoxon rank-sum tests). Median Spike-specific CD8+ T cell responses to SARS-CoV-2 WA1/2020 and B.1.617.2 (delta) were below the limit of detection in vaccinated uninfected individuals but were 2.7- and 4.4-fold higher, respectively, in vaccinated infected individuals (**Fig. 2B**; P=0.002 and P=0.002, respectively, Wilcoxon rank-sum tests). Similar trends were observed for ELISPOT responses and CD4+ and CD8+ T cell responses in individuals who received the BNT162b2 and mRNA-1273 vaccines (**Fig. S4**).

**Figure 2.**
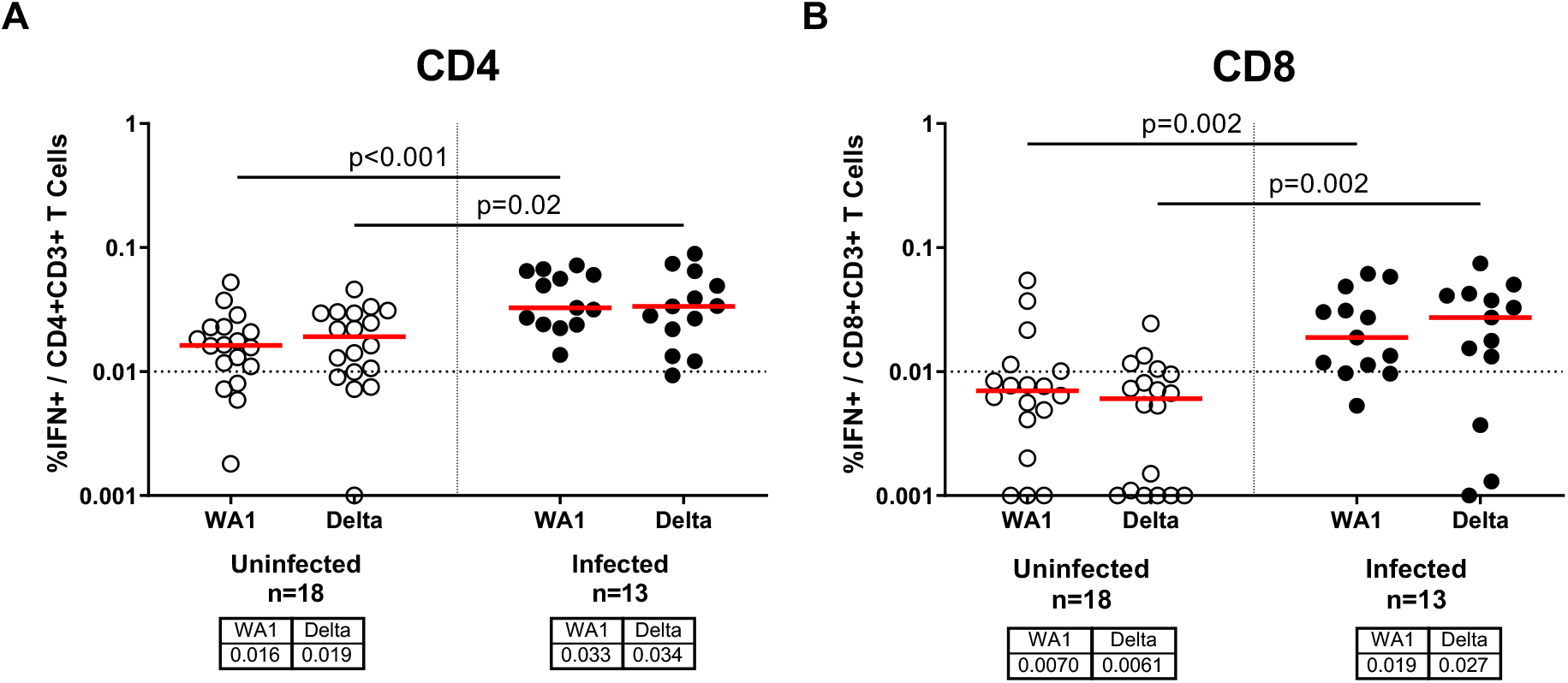
T cell responses in COVID-19 vaccinated individuals with and without confirmed SARS-CoV-2 breakthrough infection SARS-CoV-2 T cell responses were measured by intracellular cytokine staining assays to measure percent IFN-γ production in CD4 T cells (**A**) and CD8 T cells (**B**) in response to pooled Spike peptides. Vaccinated uninfected (open circles) and vaccinated infected (filled circles) individuals are shown. Assays were conducted using pooled Spike peptides from WA1/2020 and B.1.617.2 (delta). Medians (red bar) for each variant are displayed below the x-axis. Dotted line indicates limit of detection. P values reflect Wilcoxon rank-sum tests.

### Mucosal Immune Responses

Antibody responses in nasal mucosa may be critical for protection against respiratory viruses. In a subset of 18 individuals from whom nasal swabs were available, we assessed RBD-specific IgA and IgG responses in mucosal secretions extracted from nasal swabs. Nasal RBD-specific IgA responses to SARS-CoV-2 WA1/2020 and delta trended 8- and 5-fold higher, respectively, in vaccinated infected individuals compared with vaccinated uninfected individuals (**Fig. 3A**). Nasal RBD-specific IgG responses to SARS-CoV-2 WA1/2020 and delta were 19- and 21-fold higher, respectively, in vaccinated infected individuals compared with vaccinated uninfected individuals (**Fig. 3B**; both P=0.002, Wilcoxon rank-sum tests). These data show that mucosal antibody responses induced by the vaccines were low or undetectable among vaccinated uninfected participants at the timepoint tested, but were higher among vaccinated participants who tested positive for SARS-CoV-2.

**Figure 3.**
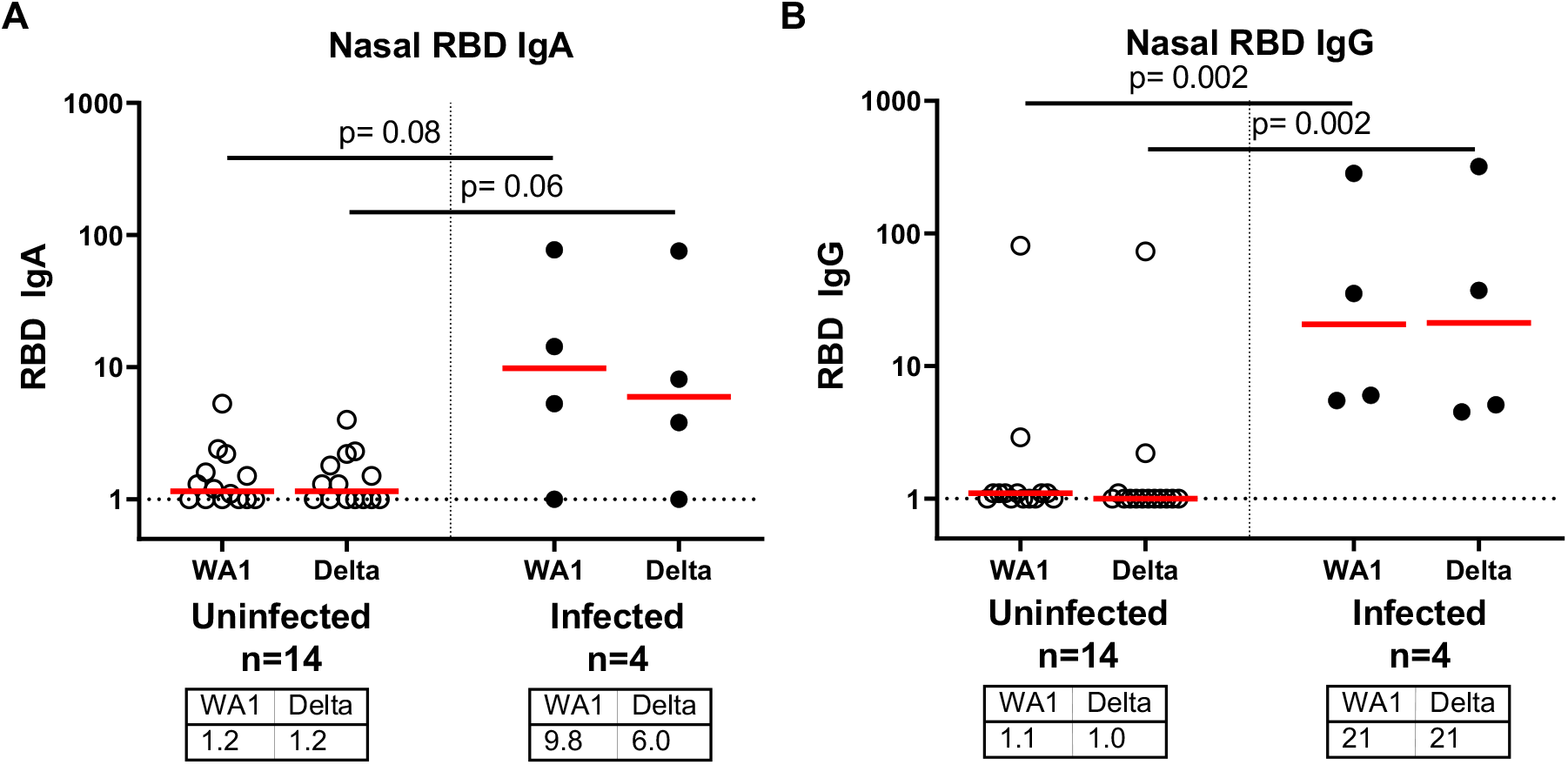
Mucosal RBD-specific nasal swab IgA and IgG responses following COVID-19 vaccination with and without breakthrough infection SARS-CoV-2 receptor binding domain (RBD)-specific IgA (**A**) and IgG (**B**) responses in nasal swabs by ELISA. Vaccinated uninfected (open circles) and vaccinated infected (filled circles) individuals are shown. Serum ELISA titers are shown against WA1/2020 and B.1.617.2 (delta) variant RBD proteins. Medians (red bar) for each variant are displayed below the x-axis. Dotted line indicates limit of detection. P values reflect Wilcoxon rank-sum tests.

## DISCUSSION

The cluster of COVID-19 infections in Provincetown, Massachusetts in July 2021 represents the first large, well-characterized outbreak of the SARS-CoV-2 delta variant in a highly vaccinated population in the United States^1^. This immunologic study provides a deeper understanding of the immune parameters associated with SARS-CoV-2 breakthrough infections in fully vaccinated individuals. Vaccinated infected individuals had markedly higher binding and neutralizing antibody responses as well as higher cellular immune responses compared with vaccinated uninfected individuals, suggesting that breakthrough infections triggered robust anamnestic immune responses. Robust and cross-reactive humoral and cellular immune responses were observed in vaccinated infected participants to all viral variants tested, including the delta variant. These data suggest important immunologic benefits of vaccination in the context of SARS-CoV-2 breakthrough infections.

Recent reports have suggested declining efficacy of the BNT162b2 vaccine after 4-6 months^7,8^, although protection against severe disease has remained high^9,10^. In the present study, participants were vaccinated a median of 5-6 months prior to the outbreak and thus were in the timeframe of declining antibody titers^11,12^. In fact, the majority of BNT162b2 vaccinated uninfected individuals had undetectable neutralizing antibody titers against the SARS-CoV-2 delta variant at the timepoint analyzed (**Fig. S1**). These immunologic parameters, coupled with the high transmissibility of the delta variant and the large community gatherings in Provincetown during and after the July 4^th^ weekend, may have contributed to the large cluster of breakthrough infections observed in this outbreak.

The robust anamnestic immune responses following breakthrough SARS-CoV-2 infections in vaccinated individuals likely contribute to the low risk of severe clinical disease in people who are fully vaccinated. Consistent with these observations are data from nonhuman primate studies in which vaccinated macaques that were not fully protected following SARS-CoV-2 challenge similarly developed high anamnestic antibody responses and rapid resolution of viral replication in the upper and lower respiratory tract^13,14^. We speculate that the extraordinarily high antibody titers observed in vaccinated individuals who develop breakthrough infections may lead to subsequent long-term protection in those individuals.

One limitation of this study is the relatively small number of individuals in this immunologic analysis compared with the large total number of individuals in this outbreak. Nevertheless, the magnitude and consistency of the immunologic differences observed between vaccinated infected and vaccinated uninfected individuals suggest the generalizability of the conclusions. Another limitation is the fact that the vaccinated infected group was generally younger than the vaccinated uninfected group, which may reflect different risk behaviors or exposures based on age. However, age did not appear to correlate with the magnitude of binding or neutralizing antibody titers in this cohort (**Fig. S2**).

In conclusion, we describe humoral and cellular immune responses in the first large, well described cluster of breakthrough infections with the SARS-CoV-2 delta variant in fully vaccinated individuals in the United States. Breakthrough infection led to a large increase in antibody and T cell responses in vaccinated individuals, suggesting important immunologic benefits of vaccination even when infection was not prevented. Moreover, anamnestic antibody responses in breakthrough infections were similar in magnitude regardless of the length of time from vaccination (**Fig. S3**), suggesting the possibility of protection against severe disease for a prolonged period of time even after serum antibody titers decline. These data provide unique insights into the immunology of breakthrough infections with the highly transmissible SARS-CoV-2 delta variant in fully vaccinated populations.

## Data Availability

All data are available in the manuscript or the supplementary material.

## ACKNOWLEDGEMENTS

The authors thank the study participants, the BIDMC Clinical Research Center, and the MA DPH. We thank Sarimer Sánchez and Julie Sklar from the Boston Public Health Commission for their assistance providing information about this immunologic study to individuals by telephone. We thank Kate Jaegle, Rachel Hemond, Esther A. Bondzie, Huahua Wan, Olivia Powers, Tianyi Ye, and Benjamin Chung from the Center for Virology and Vaccine Research for enrollment, processing, and assisting with assays for the BIDMC COVID-19 Biorepository used in this study. We thank MesoScale Discovery for providing the kits for the ECLA multiplexing kits used in this study.

## Data sharing

A.Y.C. and D.H.B. had full access to all the data in the study and takes responsibility for the integrity of the data and the accuracy of the data analysis. All data are available in the manuscript or the supplementary material. Correspondence and requests for materials should be addressed to D.H.B.

## Author Contributions

AYC: conceptualization, formal analysis, resources, investigation, data curation, writing original draft, visualization

DHB: conceptualization, formal analysis, writing original draft, review & editing, funding acquisition, supervision

CMB: conceptualization, investigation

KAM: investigation, methodology, data curation

JY: investigation, methodology, data curation

JL: investigation, methodology, data curation

CJD: resources, methodology, data curation

AC: project administration, methodology, investigation

DT: conceptualization, investigation

JLA: project administration, resources, data curation

MR: project administration, resources, data curation

DS: resources, methodology, investigation

KA: resources, methodology, investigation

RA: project administration, resources, data curation

TA: resources, methodology, investigation

SG: resources, methodology

MS: resources, methodology, investigation

LBR: resources, data curation

MRH: validation, resources, formal analysis, review & editing, supervision

LCM: conceptualization, investigation, review & editing

---

**Supplementary Figure S1.**
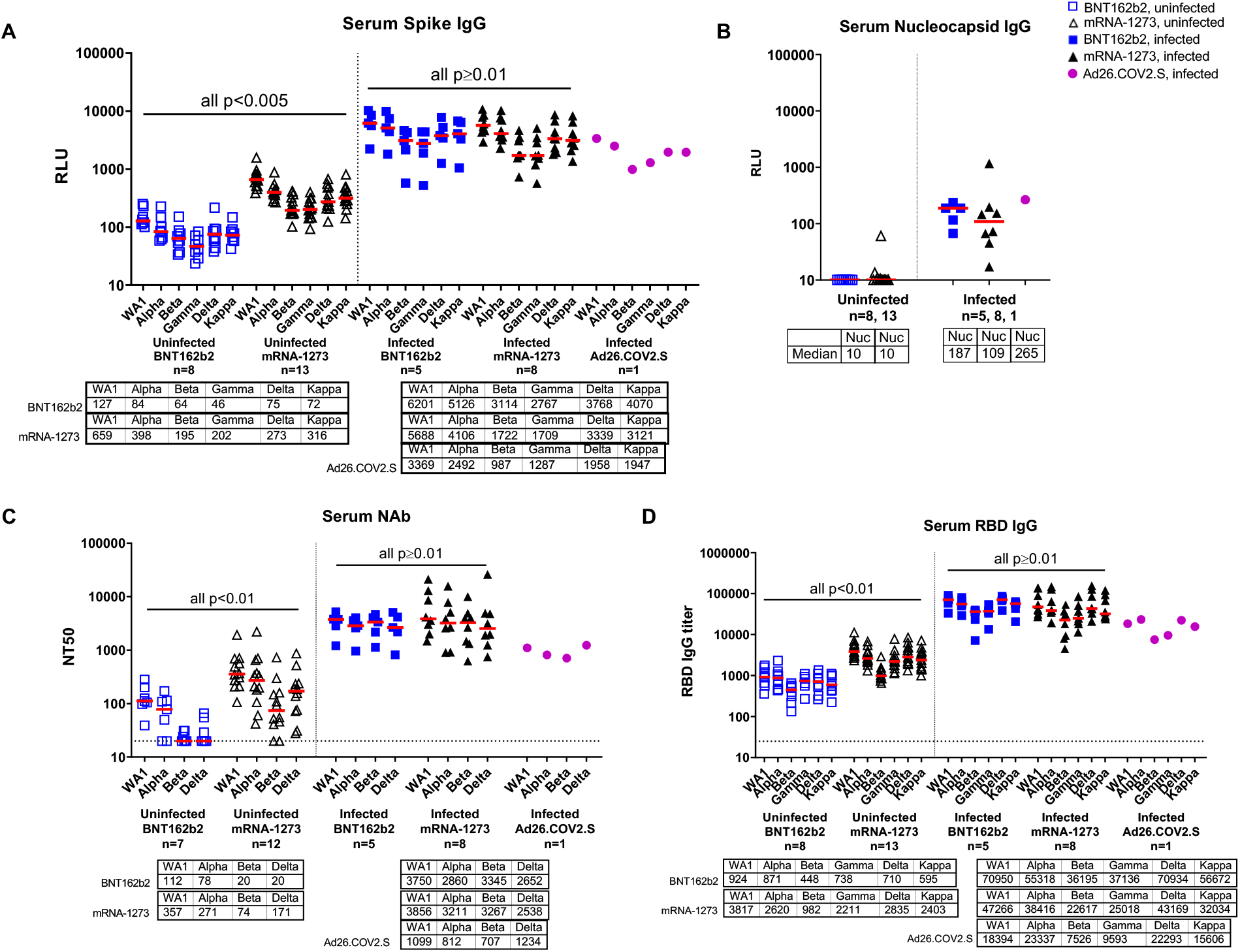
Antibody responses in COVID-19 vaccinated individuals with and without confirmed SARS-CoV-2 breakthrough infection, by vaccine type Serum antibody responses in BNT162b2 (blue), mRNA-1273 (black), and Ad26.COV2.S (magenta) vaccinated individuals as part of the SARS-CoV-2 outbreak investigation in Provincetown, Massachusetts. Vaccinated uninfected (open symbols) and vaccinated infected (filled symbols) individuals are shown. IgG binding antibody titers by multiplexed electrochemiluminescence (ECLA) assays against SARS-CoV-2 Spike from WA1/2020, B.1.1.7 (alpha), B.1.351 (beta), P.1 (gamma), B.1.617.2 (delta), and B.1.617.1 (kappa) variants in **A** and against the non-vaccine antigen Nucleocapsid (Nuc) in **B** are displayed in relative light units (Meso-Scale Discovery kits 7 and 13). **C**. Pseudovirus neutralization titers measured as 50% reduction (NT50) of luciferase expression. **D**. ELISA IgG titers to SARS-COV-2 receptor binding domain (RBD) variants. Medians (red bar) for each variant are displayed below the x-axis. Dotted line indicates limit of detection. P values (Wilcoxon rank-sum test) compare each Spike variant or Nucleocapsid between participants who received BNT162b2 and those who received mRNA-1273.

**Supplementary Figure S2.**
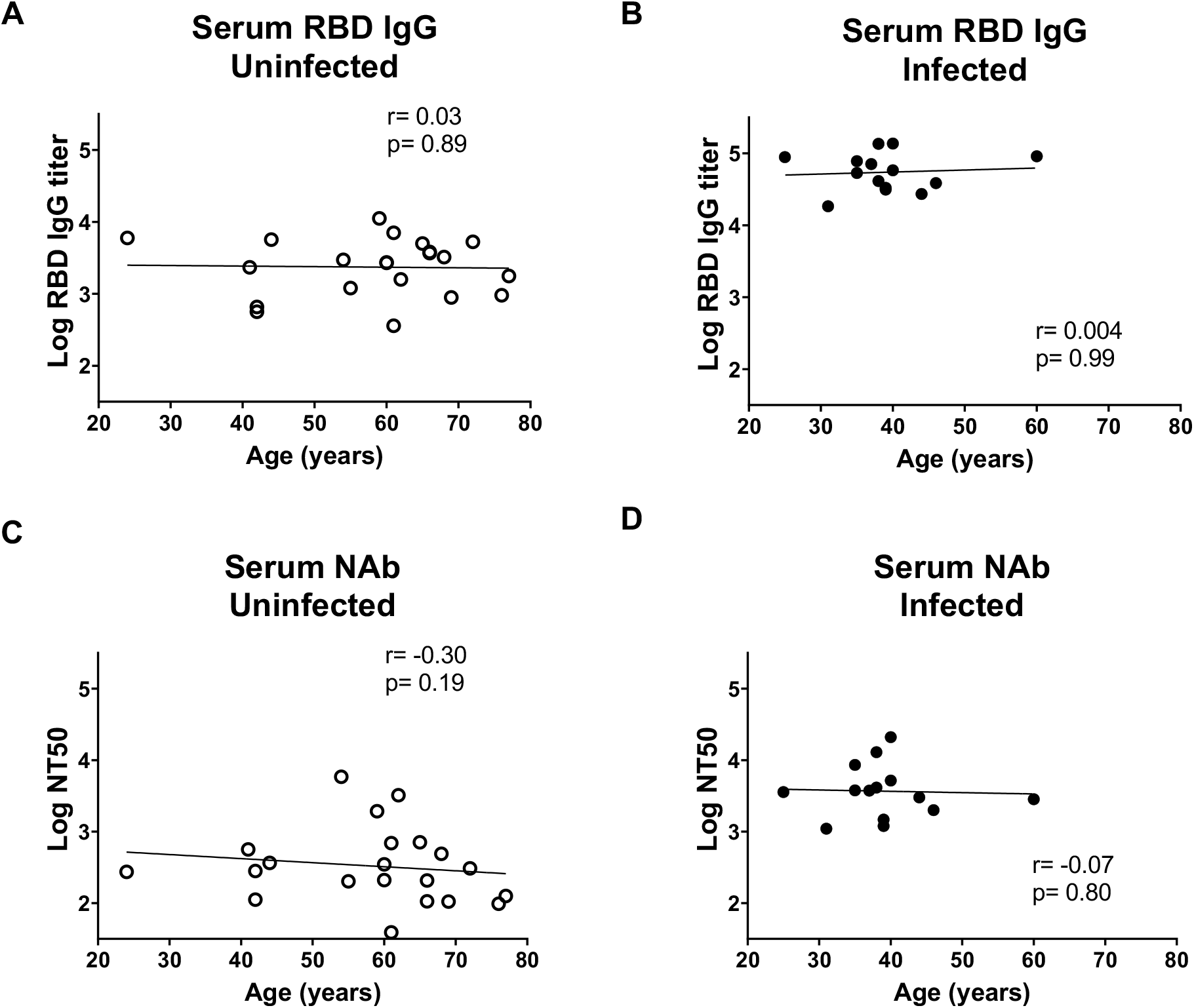
RBD-binding serum IgG titer and neutralizing antibody titer by age Scatter plot of serum RBD-specific ELISA IgG titer (**A, B**) and pseudovirus NT50 neutralizing antibody (NAb) titer (**C, D**) against WA1/2020 as a function of age. Vaccinated uninfected (**A, C**) and vaccinated infected (**B, D**) individuals are shown with line of best fit. Spearman correlation (r) and p-value are listed for each plot.

**Supplementary Figure 3.**
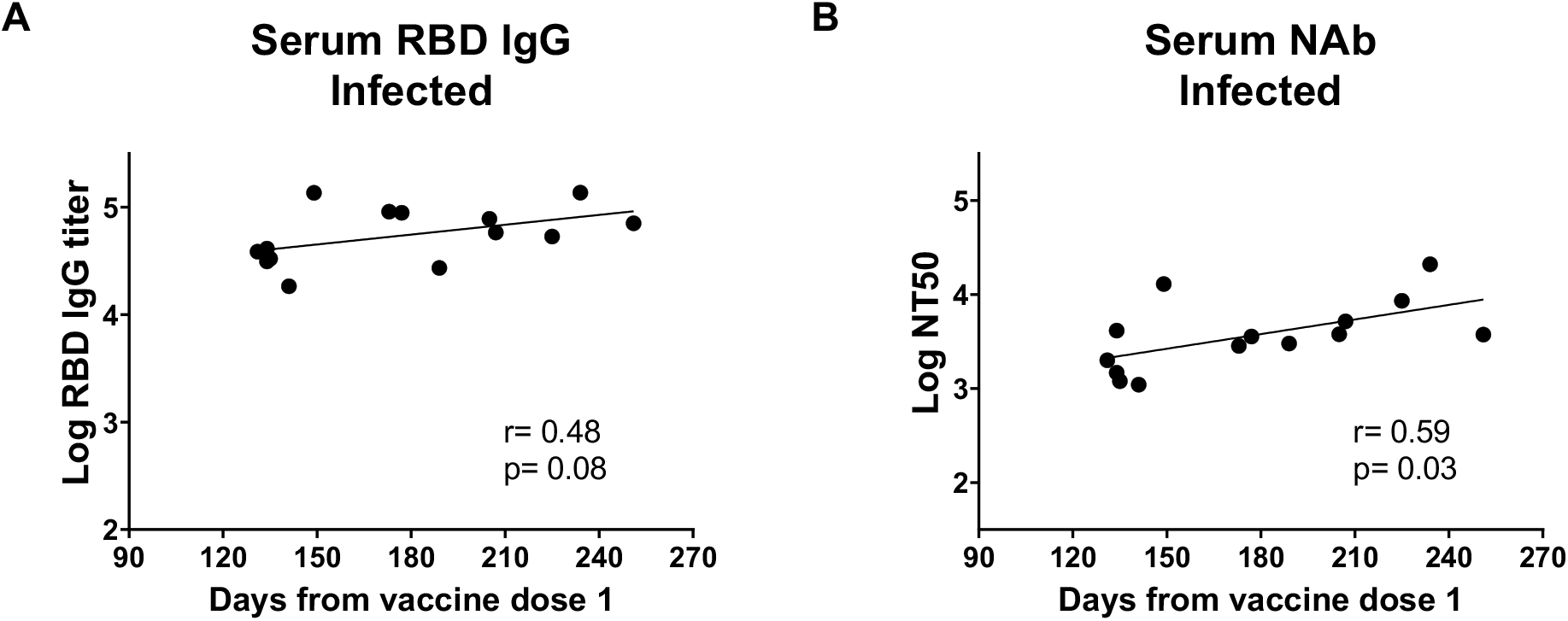
Serum RBD-binding IgG titer and neutralizing antibody in vaccinated infected individuals by days from first COVID-19 vaccine dose Scatter plot of serum RBD-specific ELISA IgG titer (**A**) and pseudovirus NT50 neutralizing antibody (NAb) titer (**B**) against WA1/2020 and days from first COVID-19 vaccine dose in vaccinated infected individuals. Data are shown with a line of best fit. Spearman correlation (r) and p-value listed for each plot.

**Supplementary Figure 4.**
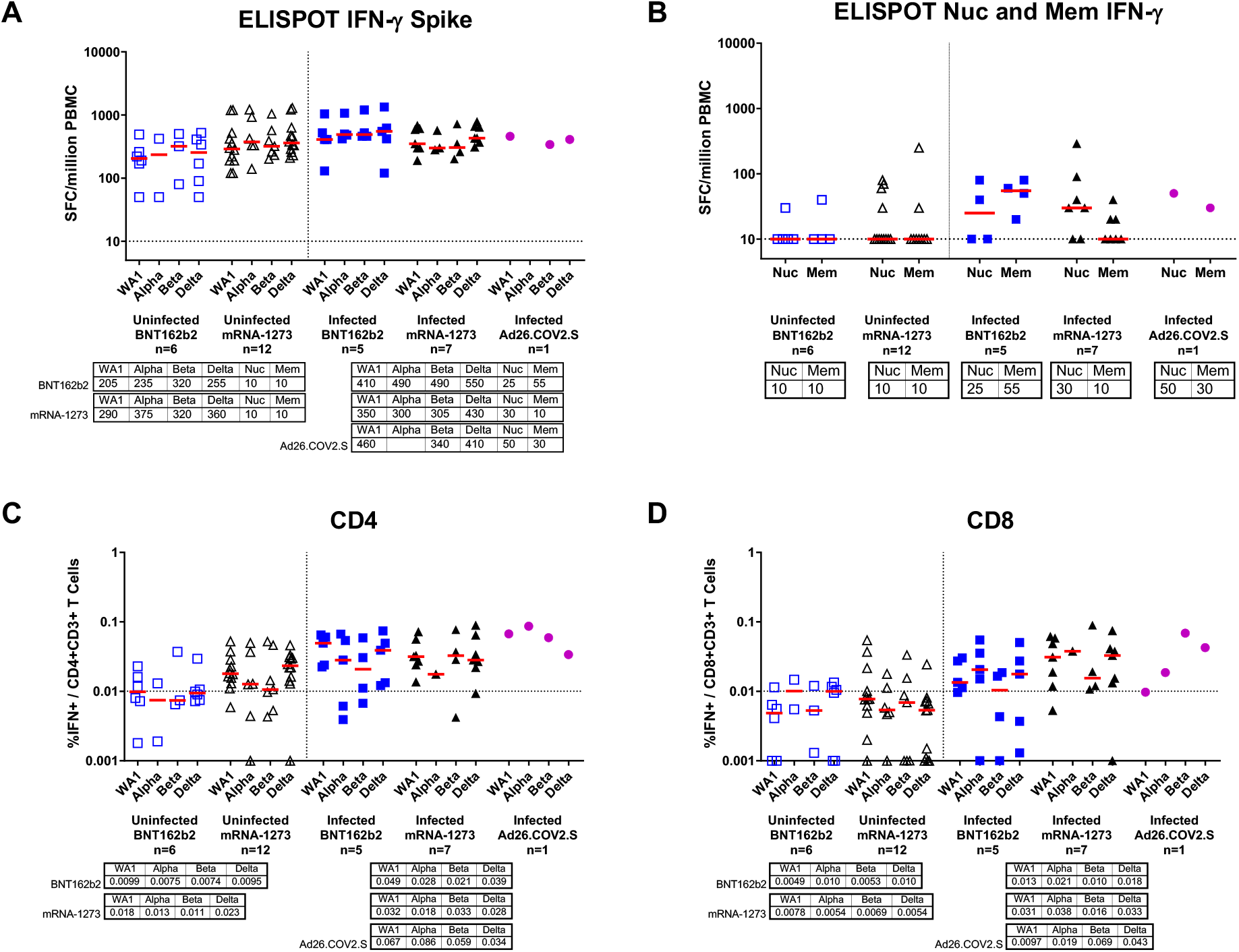
T cell responses in COVID-19 vaccinated individuals with and without confirmed SARS-CoV-2 breakthrough infection, by vaccine type SARS-CoV-2 T cell responses in BNT162b2 (blue), mRNA-1273 (black), and Ad26.COV2.S (magenta) vaccinated individuals were measured by IFN-γ ELISPOT assays after stimulation with pooled WA1/2020 Spike peptides from variants B.1.1.7 (alpha), B.1.351 (beta), and B.1.617.2 (delta) (**A**) or non-vaccine antigens Nucleocapsid (Nuc) and Membrane (Mem) (**B**). Intracellular cytokine staining (ICS) assays measured percent IFN-γ production in CD4 T cells (**C**) and CD8 T cells (**D**) in response to Spike peptides from variants. Vaccinated uninfected (open symbols) and vaccinated infected (filled symbols) individuals are shown. Medians (red bar) for each variant are displayed below the x-axis. Dotted line indicates limit of detection.

